# How 3D Printing and Social Media Tackles the PPE Shortage during Covid – 19 Pandemic

**DOI:** 10.1101/2020.04.27.20081372

**Authors:** Nick Vordos, Despoina A. Gkika, George Maliaris, Konstantinos E. Tilkeridis, Anastasia Antoniou, Dimitrios V. Bandekas, Athanasios Ch. Mitropoulos

## Abstract

During the recent Covid-19 pandemic, additive Technology and Social Media were used to tackle the shortage of Personal Protective Equipment. A literature review and a social media listening software were employed to explore the number of the users referring to specific keywords related to 3D printing and PPE. Additionally, the QALY model was recruited to highlight the importance of the PPE usage. More than 7 billion users used the keyword covid or similar in the web while mainly Twitter and Facebook were used as a world platform for PPE designs distribution through individuals and more than 100 different 3D printable PPE designs were developed.

## 1. Introduction

Coronavirus Disease 2019 (Covid-19) or SARS – CoV – 2 has already spread worldwide and affected more than 2.000.000 people, with almost 137.000 deaths, according to the Johns Hopkins University (April 16, 2020) [1]. The case fatality range for the ages between 40 – 49 estimated at 0.4% and increased at 14.8% for the 80+ age range. The epidemiological statistics were similar for most cases with some notable exceptions, such as Italy which did not follow the general rule [2]. One of the most important factors enabling the wide spread of COVID 19 is its very short diameter size, ranging between 60 and 140 nm, which renders the use of Personal Protective Equipment crucial for the protection of health professionals[3]–[6].

On October 2014, the World Health Organization (WHO) updated the PPE instructions, suggesting the use of boot covers, inner and outer gloves, masks, face protectors, surgical hood coverall, outer apron and face shield [7]. PPE is used as a shield between the health professionals and germs and must be used by the hospital staff during serious situations like a pandemic. PPE are one of the measures for the improvement of occupational health and safety [8] Special care must be taken to avoid contamination at the disposal of it [9]. PPE can generally be described as nonpharmaceutical barriers against the virus dissemination and one of the parameters accounted for in the Quality Adjusted Life Year (QALY) model [10]. The QALY allows for the combined study of outcomes of any health-related actions and their effect on mortality into a single indicator, thus establishing a method that enables comparisons across multiple disease areas.

The most significant government measure for the control of Covid -19 is social distancing. Social distancing has resulted in an increase of social media use. Social media is the modern way of communication and allows people to interact with each other via text, videos, pictures and music. More than 40% of the world’s population is a social media user and more than 1.5 billion are daily active users [11]. One of most impressive use of social media is for health - related purposes [12]. A large part of the population is informed for health issues and communicate with health professionals through social media [13]. The best advantage of that form of contact is that it is a means of mass communication and offers the ability to facilitate disease surveillance [12], [14].

At the same time, medical applications for 3D printing are expanding rapidly and are expected to revolutionize health care [15]. The first 3D printers were invented from Hideo Kodama and Charles Hulls in the early 1980s [16]. Additive technology used in various sections such as the Aerospace and Automotive industry, military, Sports field, architecture, toys industry and bioengineering with different benefits and disadvantages [17]. Since then, different 3D printing methods have been used, based on extrusion, powder solidification and liquid solidification, with different types of materials as building materials [18].

The aim of the present article is to explore the relationship between social media and 3D printing, in the context of the recent Covid-19 pandemic. We will analyze which types of Personal Protective Equipment can be printed, and how the 3D printing users can be coordinated to achieve mass printing volumes.

## 2. Methods and Materials

### 2.1 Overview

Two independent searches have been conducted, in order to examine the degree that social media has affected the development and dissemination of PPE and medical equipment parts. They explore how 3D printed designs were utilized to address the Covid-19 pandemic, as well how the QALY model can be applied in this case to measure the effects of the use of PPE. In the first in depth search, social media was studied using targeted keywords, while an official database was implemented and the QALY model was applied.

### 2.2 Social Media and 3D printing of PPE

Since Covid – 19 appeared in the Wuhan district in China, several drawings of PPE were shared between social media users, 3D community and individual citizens. A specialized social media software (Awario) was recruited for gathering information about content which refers to Covid-19, 3D printing and PPE. Targeted queries were performed on popular social media such as Facebook, Twitter, Instagram, YouTube, Reddit and also on News/Blogs and the Web in general. The searches were conducted between January 1, 2020 through April 14, 2020. The results were further examined for details on the representative designs of PPEs.

The search strategy consisted of using two generic lists of terms with one specific keyword. The first keyword set refers to the different names of the novel coronavirus and will be referred to as Keywords Set 1 (KS1), with the following syntax:

“Covid OR Covid 19 OR Coronavirus OR Covid_19 OR Covid-19 OR SARS-CoV-2”,

The general Keywords Set 2 (KS2) comprised of terms which correlate to 3D printing technology and have the following Boolean syntax:

“Design” OR “3D” OR “Additive Technology” OR “3D printing” OR “Print”

The specific keyword is related to the PPEs and specific medical equipment, which for this study are “PPE”, “Personal Protective Equipment”, “Face Shield”, “Goggles”, “Gloves”, “Boots”, “Surgical Hood”, “Valves”, “Ventilator” and “Respirator”.

The syntax of the queries that were submitted in the social media listening software has the following format:

“KS1” AND “KS2” AND “X”,

where X is one of the specific keywords mentioned. For example, when looking into which social media post includes the keywords Covid (or similar)” and 3D printing (or similar) and PPE, the following query syntax “KS1” AND “KS2” AND “PPE” was used.

### 2.3. PPE and QALY

There are two major parameters that have been studied, the life expectancy of health professionals and the average life expectancy until the death of someone who became ill with Covid - 19 which suggests that hygiene rules or Personal Protective Equipment have not been used or are being misused. Data on life expectancy were obtained from WHO and OECD, while the calculation of the average lifespan was researched in the scientific databases PubMed and Scopus with the keywords “Symptoms” AND “Death” AND “Covid” AND “Days” [19], [20]. The results of the search were analyzed and summarized in the next section.

## 3. Results

Table 1 provides the results from the social media listening software. The first three columns are comprised by the aforementioned Keyword sets used in the query, the columns 4 –10 are the social media and web sources, while column 11 contains the result of how many users in the social media have impacted the query. For example, for the query Face Shield AND KS1 AND KS2, 3.1 Million users reacted (like, share, post, etc.) in the Social Media. The first line presents the results for the query of KS1 which refers to the different expressions of the novel coronavirus. More than 7 billion users reacted in the social media, web, blogs and news website for the mentioned timeframe. 3.9 million and 4.2 million users reacted when using the KS1, KS2 and the keywords PPE or “Personal Protective Equipment” respectively. For each PPE the number of the users varied between 50.6K to 9.1M, while for valves and ventilators the numbers ranged between 1.4 M and 3.1M. The bolded percentages indicate the highest percentages in Social Media, Blogs or Web appearance on listening results.

**Table 1.** Social Media listening Results. *B: Billion, M: Million, K: Kilo

| Keywords |  |  | Twitter | Facebook | Instagram | YouTube | Reddit | New/Blogs | Web | Total (Reach)* |
| --- | --- | --- | --- | --- | --- | --- | --- | --- | --- | --- |
| KS1 |  |  | 14,5% | **63,0%** | 8,7% | 8,7% | 0,5% | 1,1% | 3,4% | 7,2 B |
| KS1 | KS2 | PPE | 12,9% | 2,4% | 0,0% | 14,7% | 1,2% | **38,8%** | 30,0% | 3,9 M |
| Covid |  | Personal |  |  |  |  |  |  |  |  |
| OR |  | Protective |  |  |  |  |  |  |  |  |
| Covid 19 |  | Equipment | 5,3% | 3,5% | 0,0% | 10,5% | 1,8% | **50,0%** | 28,9% | 4,2 M |
| OR | Additive | Face Shield | 6,6% | 2,0% | 0,0% | 21,7% | 0,0% | **42,8%** | 27,0% | 3,1 M |
| Coronavirus | Technology | Goggles | 0,0% | 0,0% | 0,0% | 0,0% | 4,0% | **64,0%** | 34,0% | 59,3 K |
| OR | OR | Gloves | 0,0% | 0,0% | 0,0% | 0,0% | 0,0% | **58,1%** | 41,9% | 126,3 K |
| Covid_19 | 3D printing | Boots | 0,0% | 0,0% | 0,0% | 0,0% | **64,7%** | 23,5% | 11,8% | 234,9 K |
| OR | OR | Surgical Hood | 0,0% | 0,0% | 0,0% | 0,0% | 0,0% | 36,4% | **63,6%** | 50,6 K |
| Covid-19 | 3D print | Valves | 4,2% | 1,4% | 0,0% | 8,5% | 0,0% | **57,7%** | 28,2% | 1,4 M |
| OR |  | Ventilator | 17,4% | 3,3% | 0,0% | 2,2% | 6,5% | **43,5%** | 27,2% | 3,1 M |
| SARS- |  |  |  |  |  |  |  |  |  |  |
| CoV-2 |  | Respirator | 9,2% | 1,9% | 0,8% | 13,0% | 5,7% | **46,2%** | 23,2% | 9,1 M |

\*B: Billion, M: Million, K: Kilo

Fig. 1 illustrates the results described in Table 1. Goggles, Gloves, boots and surgical hoods have the lowest resonance in social media and web, in contrast to respirators and face shields (9100 K and 3100K users respectively used combinations of keywords). Ventilators and valves are also prominent in the results. Fig 2 is a graphical representation of table 1 results where each combination of keywords is presented in relation to the percentage of appearances in the corresponding social media or web. In many cases, zero results were returned, while in most of the cases with high percentages, those were found through websites and blogs.

**Figure 1.**
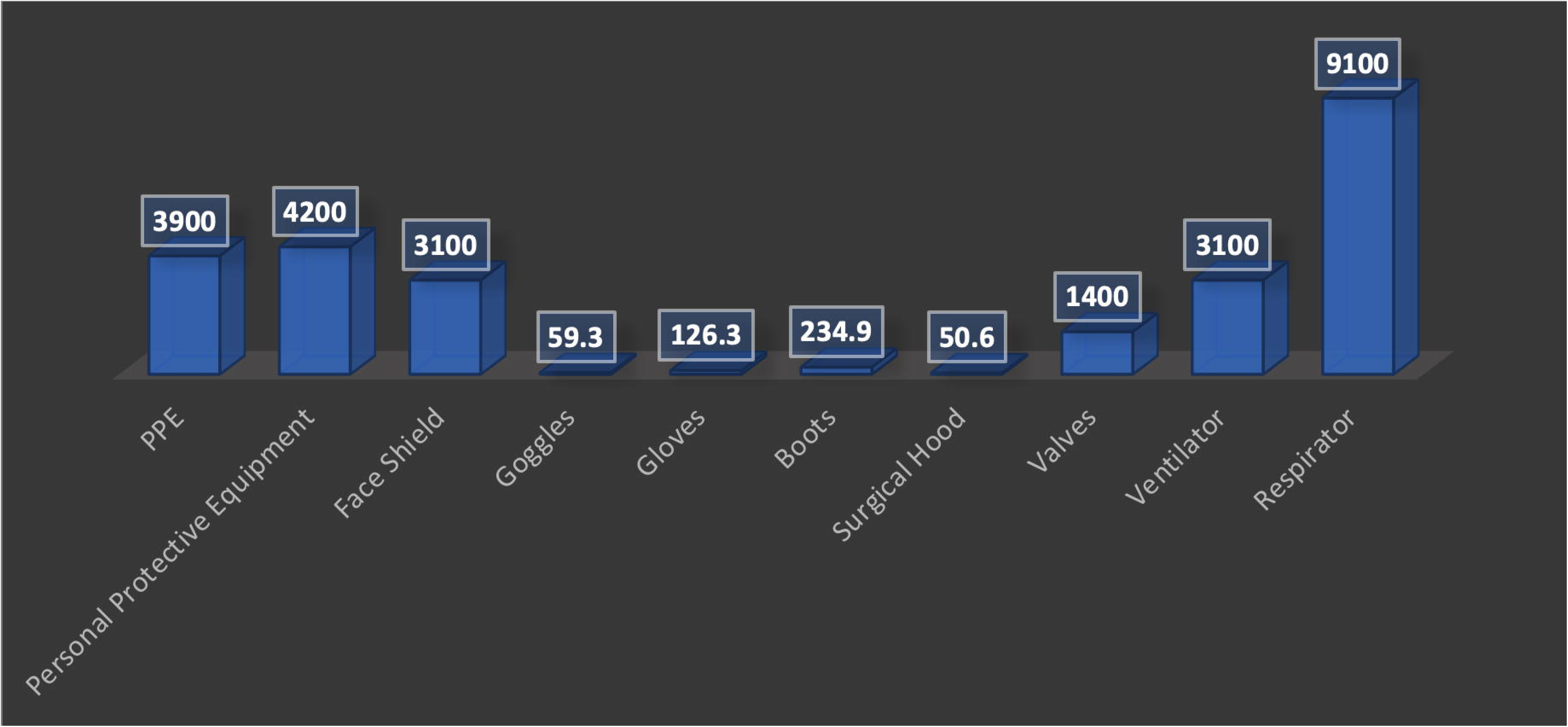

**Figure 2.**
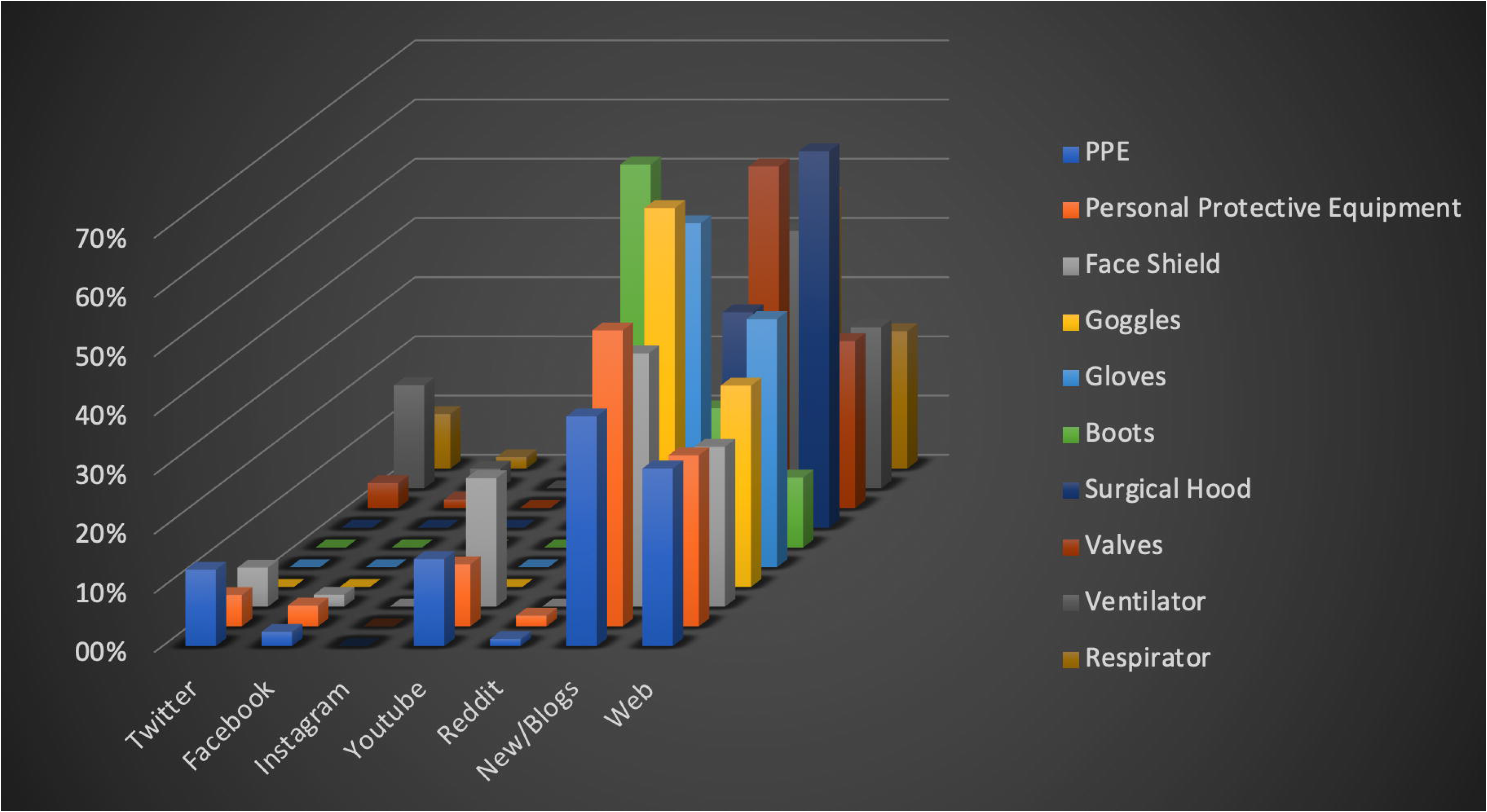

Table 2 shows which of the WHO proposed PP equipment are printable with personal or university laboratory 3D printers. The third column of the table shows whether they can be sterilized while the fourth column shows the number of the different designs appeared in Social Media. 109 different designs of face masks (similar in many cases) were proposed in order to help the health workers avoid exposure. More than 30 designs were proposed for respirators, two designs for goggles, while there were not any design for boot covers.

**Table 2.** PPE which can be 3D printed or not and the number of designs.

| **PPE** | **3D Print** | **Sterilized** | **Number of Designs** |
| --- | --- | --- | --- |
| Boot Covers | Yes (Reusable) | Yes | 0 |
| Gloves | No | - | 0 |
| Masks | No | - | 0 |
| Respirator | Yes (Reusable) | Yes | 36 |
| Surgical Hood | No | - | 0 |
| Outer Apron | No | - | 0 |
| Medical Glasses /Goggles | Yes (Reusable) | Yes | 2 |
| Face Shields | Yes (Reusable) | Yes | 109 |

Based on our search, more than 140 Social Media Groups from 36 countries and more than 18000 individual members participated in the 3D printing effort for Covid 19. The main ideas for helping to fight the novel coronavirus except the previously mentioned PPE, refer to Oxygen venturi valves, Hands free door handle attachments, Printable face masks, Mask adjuster, Ventilators, Test swabs, Valve for Scuba masks, Spirometry Sensors, Ventilator Splitter and Oxygen Filter Housings. Representative PPE designs are illustrated in Fig. 3.

**Figure 3.**
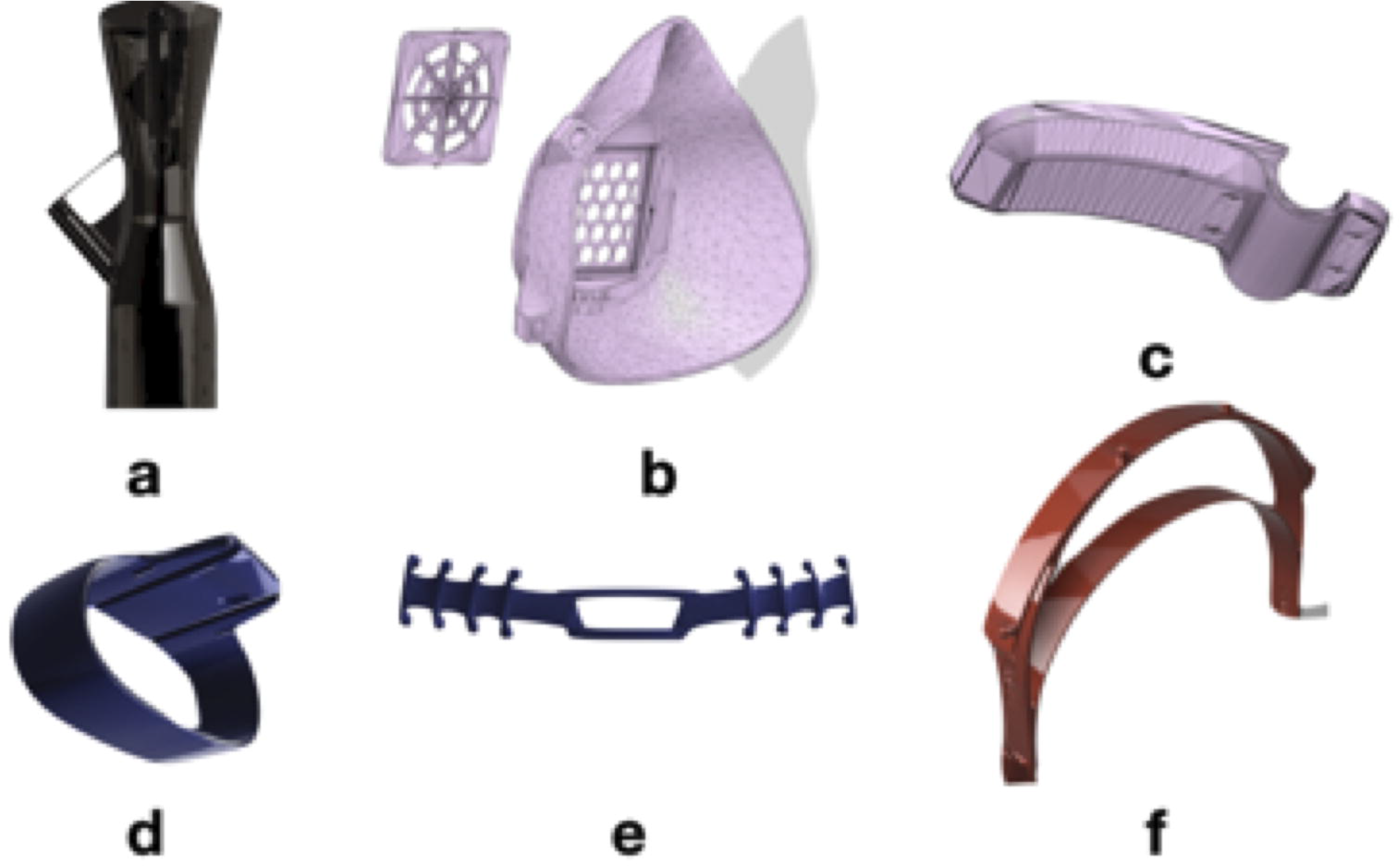

According to the World Health Organization, life expectancy on average in the world is 72 years. A more detailed study of the Organization for Economic Co-Operation and Development (OECD) showed that life expectancy for men is 77.7 years while for women it is 83.1 years. By the time of this study, more than 200 healthcare professionals have died from the novel coronavirus disease [21]. 29% were physicians in the USA, with ages ranging between 56 - 65 years old and 17% were over 66 years old, while in Europe 38% of healthcare professionals were older than 55 years old [22], [23].

The search in the scientific databases for the time interval between the onset of the symptoms and the death showed, as expected, different results. Table 3 shows the results from the Scopus bibliographic database, which procured 17 articles, as opposed to 12 from PubMed.

**Table 3.** Scopus and PubMed results for duration between symptoms and death

| **Keywords** | **PubMed** | **Scopus** | **Days** | **Ref** |
| --- | --- | --- | --- | --- |
| Days | 12 | 17 | 8 | [23] |
| AND |  |  | 11,5-20 | [24] |
| Symptoms |  |  | 16 | [25] |
| AND |  |  | 17,8 | [26] |
| Death |  |  | 23 | [27] |
| AND Covid |  |  | 23,4 | [28] |

The total number of those results that included the desired information was 6, whereas the time between symptoms and death appeared between 8 and 23.4 days.

Figure 4 depicts the QALY concept and illustrates potential gains if a subject uses a PPE as opposed to not using one. The area between the curves corresponds to the total QALY gains by using a PPE.

**Figure 4.**
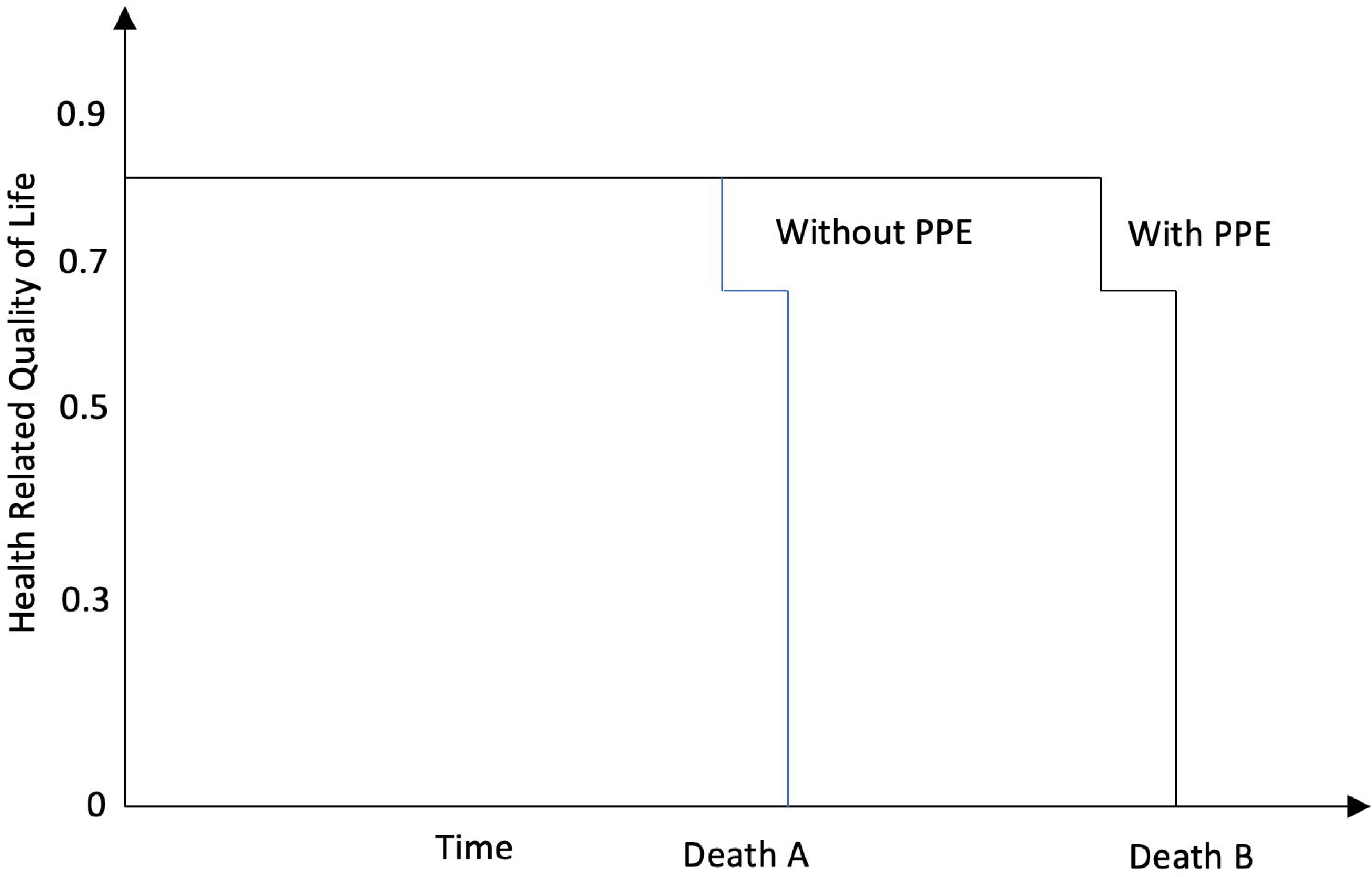

The lower curve depicts the scenario of no use of face shield, up to the time of the subject’s death (Death A). Similarly, the top curve depicts the path if a face shield has been used, also till the time of the person’s death (Death B); the Health-Related Quality of Life in this case persists at higher levels for much longer.

## 4. Discussion

In this study, the relationship between the epidemic, the social media and the use of three-dimensional printing is presented. The ultimate goal of the article is to highlight the use of Social Media and additive technology in general from common users, to address the lack of basic personal protective equipment or even parts of machinery used by health professionals. The QALY model was employed to illustrate the necessity of PPE use.

More than 5000 scientific articles, for the role of social media during different crisis types (terrorist attacks, hurricanes, tsunamis, earthquakes, etc.) appear in scientific databases (Scopus, PubMed and Google Scholar). In many cases, social media can operate as a crisis platform to generate community crisis maps [24], [25]. There are four types of connection between the users of social media during a crisis: i) the authorities communicate with citizens (statements, information, instructions, etc.), ii) authorities communicate with other authorities for an inter-organizational crisis management, iii) citizens to citizens (photographs, information, communities) and iv) the citizens communicate with authorities [26].

As was expected, social distancing due to Covid – 19 resulted in an increase in social media use. Only in March 11, 2020, more than 19 million mentions related to the new coronavirus. In general, social media are used for communication between citizens or simply to kill time and to reduce distancing. On the other hand, healthcare organizations like WHO used social media to provide information and rules of hygiene to battle Covid – 19. Almost all platforms set at the main page a joint statement for the fight of the pandemic [27].

The spread of Covid – 19 in the global community, in a short period of time, has resulted in the rapid depletion of PPE. Risk to health or the danger of accident of professional health workers can be minimized with the use of PPE [28]. The choice of the appropriate PPE must be done according to the degree of exposure in gems, the material of PPE and the ergonomics of it [29]. Many healthcare professionals, from different countries use social media to express the lack of respirators, gowns and face masks [30]–[32]. WHO also uses social media to ask from government and industry to increase the manufacturing process of PPE by 40% [33].

The interaction between authorities and other authorities, as well as authorities and citizens and vice versa was as expected. The reaction was impressive between the citizens, who used social media to tackle the shortage of PPE and to help the doctors and nurses during the pandemic. Worldwide, social media groups exchanged ideas, drawings, schematics and technical instructions, in order to produce face shields, reusable respirators, ventilators and other PPE [34], One of the most important actions of social media users is the use of 3D printing technology to produce specific PPE. Social networks have the potential to combine both information from the official sources and popular information from citizens in a short period of time, making them a valuable tool in crisis management [35]. Previous studies show that in times of crisis, citizens seem to be more cooperative and better able to respond to authorities’ instructions via social media [36]. The main advantage of using these tools, is the dynamic capability they provide to disseminate the information [37], Unfortunately, the use of media has not only advantages, but also disadvantages, such as unreliability of the information, and inefficiency and dispersal of panic [13], [38]. At the time of the Covid – 19 pandemic, the network platform use rose by at least 20%, communicating information for the virus spread and for the shortages of PPE [39]. This information has motivated the global community to look for possible solutions to these problems. The formulation of relevant groups was immediate, and designs for the possible solutions were placed in digital repositories, where the main technology used is 3D printing.

It is not the first time that 3D printing and additive technology in general has been used to provide solutions in the field of medicine and biomechanics. Various technologies and materials have been used to print surgical instruments, dentures, organ dummies, etc.[18]. Only in the United Stated of America, there is an estimated amount of 444.000 printers, then the United Kingdom has more than 168.000 printers, while Germany occupies the 3^rd^ place in the world [40], [41]. It is the first time that the community of 3d printer home users has come together and mass-produced PPE. More than 180.000 users worldwide can produce up to 6 face shields in 10 hours each, depending on the capabilities of printers and the design. Assuming that a country like Greece has about 500 printers, then in one day more than 6.000 face shields can be produced, enough to equip doctors, nurses, rescuers, staff working with patients in general, in a short period of time.

In our study, the queries that included the general terms PPE and Personal Protective Equipment show the most results in user response, as expected, because they concern general fields and not individual PPE. In most cases, the largest percentage is due to the impact of social media users on news / blogs and WEB, due to the fact that social media groups display their products/results on news websites. Proof that the teams were organized through social media is that in the PPEs that can be printed (Face Shields, Respirator but also Valves and ventilators) show increased percentages on Twitter, Facebook and YouTube, while in the rest they are zero.

Although several designs have been developed for 3D printing, the biggest concern is whether all of them can be used. Some of these designs have been approved by NIH and FDA. Particular attention should be paid to the materials used for their production, but also to the way they are used so as not to endanger human lives. The American Society of Anesthesiologists made a statement about the possibility of having multiple patients per ventilator. Characteristically, they state a number of reasons why it is impossible to do so, for example, “Volumes would go to the most compliant lung segments. Positive end - expiratory pressure, which is of critical importance in these patients, would be impossible to manage.” [42], For such types of reasons, even after about three months from the beginning of the pandemic, licenses for the manufacture of respirators by individuals in the vast majority, have not been issued

In figure 5 a proposed process used before 3D printing is illustrated. The need for PPE and medical supplies triggers the beginning of the process. Individuals are usually searching for designs and solutions for the problems concerning the local community through social media. Accordingly, they should verify whether the final product has been approved by the competent authorities. In case the proposed design lacks a license, a similar design that has the relevant license should be selected. Following, the prototype is printed and inspected from the local healthcare professionals for micro adjustment, if necessary. Finally, if all the previous steps are completed, the product can be printed in mass quantities.

**Figure 5.**
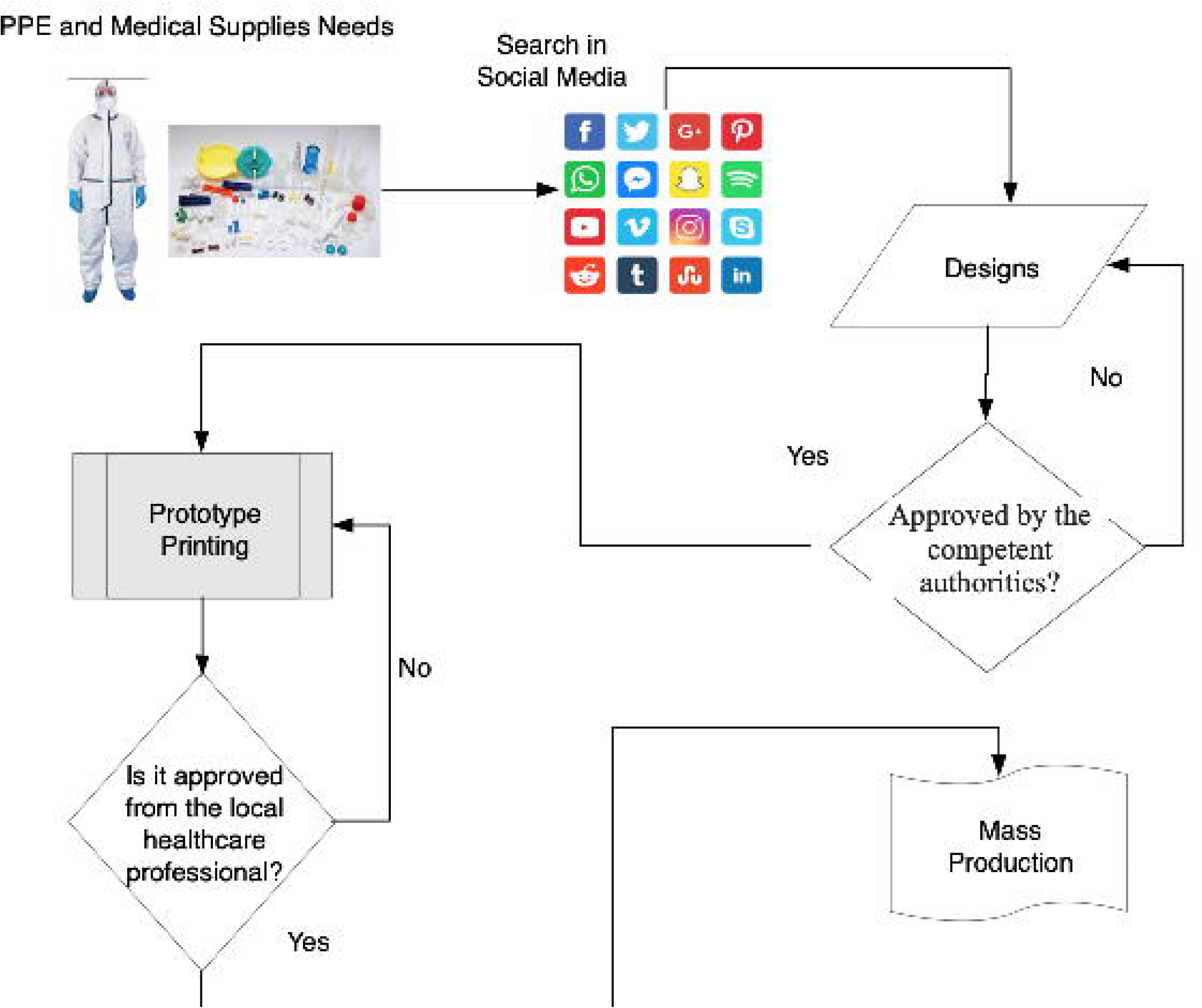

The QALY allows for the combined study of outcomes of any health-related actions and their effect on mortality into a single indicator, thus establishing a method that enables comparisons across multiple disease areas. Throughout their life, people have different health states, which are weighted based on the preassigned utility ranks. The QALY model was applied as shown in figure 4, taking into account that one of the main reasons of viral infection affecting the respiratory system is the non-usage or poor application of protective equipment as well as the fact that health professionals are more exposed and subsequently more likely to get infected. Patients using PPE gained quality of life compared to health worker professionals from Europe or US, without the usage of PPEs.

## 5. Conclusion

This study examined the way individual social media users and 3D printer owners tackle the PPE shortage during a pandemic. Social media influences the problem in multiple levels: Firstly, they highlight the problem, in this context the lack of PPEs, secondly they encourage and promote the formation of task forces/teams from the general population with a relevant interest, thirdly they provide the means for exchanging information and technology and finally they can identify the number of required 3D printers in a local, national, or even international level needed to achieve the task.

At the same time the role of PPE as necessary equipment for health professionals is fundamental. The QALY model was employed to show the importance and effect of using personal equipment on life quality and expectancy. During the recent coronavirus pandemic, the world faced a serious shortage of PPE. Individuals and Universities co-ordinated their action using web networking and social media in order to produce around 150 3D printable designs of PPE that got developed and distributed. Social networking and 3D printing combined can be seen as a new tool for tackling pandemic emergency situations.

## Data Availability

Non-digital data available

